# Higher hospitalization and mortality rates among SARS-CoV-2 infected Persons in Rural America

**DOI:** 10.1101/2021.10.05.21264543

**Authors:** Alfred Jerrod Anzalone, Ronald Horswell, Brian Hendricks, San Chu, William Hillegass, William Beasley, Jeremy Harper, Clifford Rosen, Lucio Miele, James McClay, Susan L Santangelo, Sally Hodder

## Abstract

**IMPORTANCE:** Rural communities are among the most underserved and resource-scarce populations in the United States (US), yet there are limited data on COVID-19 mortality in rural America. Furthermore, rural data are rarely centralized, precluding comparability across urban and rural regions.

**OBJECTIVE:** The purpose of this study is to assess hospitalization rates and all-cause inpatient mortality among persons with definitive COVID-19 diagnoses residing in rural and urban areas.

**DESIGN, SETTINGS, AND PARTICIPANTS:** This retrospective cohort study from the National COVID Cohort Collaborative (N3C) examines a cohort of 573,018 patients from 27 US hospital systems presenting with SARS-CoV-2 infection between January 2020 and March 2021, of whom 117,897 were hospitalized. A sample of 450,725 hospitalized persons without COVID-19 diagnoses was identified for comparison.

**EXPOSURES:** ZIP Codes provided by source hospital systems were classified by urban-rural gradient through a crosswalk to the US Department of Agriculture Rural-Urban Commuting Area Codes.

**MAIN OUTCOMES AND MEASURES:** Primary outcomes were hospitalization and all-cause mortality among hospitalized patients. Kaplan-Meier analysis and mixed effects logistic regression were used to estimate 30-day survival in hospitalized patients and associations between rurality, hospitalization, and inpatient mortality while controlling for major risk factors.

**RESULTS:** Rural patients were more likely to be older, white, have higher body mass index, and diagnosed with SARS-CoV-2 later in the pandemic compared with their urban counterparts. Rural compared with urban inhabitants had higher rates of hospitalization (23% vs. 19%) and all-cause mortality among hospitalized patients (16% vs. 11%). After adjustment for demographic and baseline differences, rural residents (both urban adjacent and non-adjacent) with COVID-19 were more likely to be hospitalized (Adjusted Odds Ratio (AOR) 1.41, 95% Confidence Interval (CI), 1.37-1.45 and AOR 1.42, CI 1.35-1.50) and to die or be transferred to hospice (AOR 1.62, CI 1.30-1.49 and 1.38, CI 1.30-1.49), respectively. Similar differences in mortality were noted for hospitalized patients without SARS-CoV-2 infection.

**CONCLUSIONS:** Hospitalization and inpatient mortality are higher among rural compared with urban persons with COVID-19, even after adjusting for several factors, including age and comorbidities. Further research is needed to understand the factors that drive health disparities in rural populations.

**KEY POINTS:** *QUESTION:* Do mortality and hospitalization rates among rural SARS-CoV-2 infected patients in the United States differ from that of their urban counterparts?

*FINDINGS:* Hospitalization and mortality rates were significantly greater among rural compared with urban patients, even after adjusting for rural-urban differences including age and comorbidities. Similarly, in persons without SARS-CoV-2 infection, mortality rates were also higher among rural patients.

*MEANING:* These findings demonstrate a pervasive rural-urban discrepancy in all-cause inpatient mortality among rural individuals in the U.S. infected with SARS-COV-2 as well as among those who were uninfected. Further research is needed to understand factors driving health disparities among rural populations.

## Introduction

The novel coronavirus (SARS-CoV-2) was the third leading cause of death in the United States (US) in 2020^1^ and is responsible for more than 650,000 deaths to date.^2^ During the initial four months of the US SARS-CoV-2 epidemic, cases were most concentrated in urban areas. However, by late 2020, rural communities experienced a surge in SARS-CoV-2 infections with some of the highest case rates in the nation.^3^ Nonmetropolitan areas constitute 97% of the US land area, with approximately 20% of the population residing in rural areas.^4^ Rural compared with urban inhabitants are older, less likely to engage in behaviors to prevent SARS-CoV-2 infection,^5^ and have a higher prevalence of comorbidities (e.g., obesity) associated with more severe COVID-19 (C19) and death.^6^ They have also experienced a greater disparity in over life expectancy over the last fifty years,^7^ which is likely linked to reduction rural providers^8^ and services that necessitate an increase in transfers to distant acute care facilities.^9^ Rural residents also face various social, economic, and environmental factors distinct from urban communities.^10^

To better understand potential drivers of C19 outcomes in rural America, we assessed hospitalization and mortality using the National COVID Cohort Collaborative (N3C), a National Institutes of Health-supported data enclave containing electronic health record information on more than 6 million persons tested for SARS-CoV-2 across 55 US sites and more than 2 million patients with a definitive diagnosis of SARS-CoV-2 infection. To our knowledge, this is the largest cohort of C19 cases in North America using data at the patient-level. Previous large-scale studies have been restricted to single states^11^ or utilized public health reporting systems.^12^ While the relationship between rural and urban hospitalization and mortality has been studied for chronic conditions, limited research has evaluated SARS-CoV-2 infected rural-urban discrepancies.

## Methods

This retrospective cohort study received Institutional Review Board approval from each investigator’s institution and was reviewed and approved by the N3C Data Access Committee. Our study cohort includes patients diagnosed between January 1, 2020, and March 31, 2021, and a demographically matched (2:1 SARS-CoV-2 uninfected-to-infected matching done at the site level as part of the N3C ingestion process based on age, gender, race, and ethnicity)^13^ comparison group of SARS-CoV-2 uninfected persons. This study followed the Enhancing the Quality and Transparency of Health Research (EQUATOR) reporting guidelines, Reporting of Studies Conducted Using Observational Routinely Collected Health Data (RECORD).^14^

### N3C Data Enclave

N3C has broad inclusion criteria, harmonizing data from 55 sites across the US.^15^ N3C collects longitudinal Electronic Health Record (EHR) or Health Information Exchange (HIE) data (with a 2-year “lookback” period to January 1, 2018) on all patients with a C19 diagnostic code (22% of all patients) or a positive SARS-CoV-2 polymerase chain reaction (PCR) or antigen test (78% of all patients) as well as uninfected patients serving as controls. Source system C19 testing protocols are mapped to standard terminologies for labs (LOINC) and conditions (ICD-10 CM and SNOMED CT) by the N3C Data Ingestion and Harmonization Workstream, which maintains a computable phenotype for defining presence of C19.^16^ To capture patients during the early stages of the pandemic (before 5/1/2020), patients with two weak diagnostic codes (such as ICD10 J80* Acute Respiratory Distress Syndrome and R43.0 Anosmia) probabilistic of C19 are also included.^17^

### Cohort Identification

Rural and urban categories were identified by 5-digit ZIP Codes that were then mapped to the 2010 Rural-Urban Continuum Codes (RUCA), distinguishing by population density, degree of urbanization, and adjacency to metropolitan areas.^18^ For purposes of this study, three categories are defined: urban, urban-adjacent rural (UAR), and nonurban-adjacent rural (NAR).^19, 20^ This classification has been commonly used to attribute rurality based on census tract or ZIP Code.^21, 22^ To validate representativeness of the cohort population with the overall US population, we compared the population percentages for each category in N3C with the US population using public datasets.^2,23^

N3C data partners are contributing institutions that encompass multiple providers and potentially multiple care sites. We developed and utilized a data robustness screening matrix to determine minimum fact reporting per patient across key domains for each data partner. This follows a similar approach used by the four source data models that all rely on data quality dashboards to enhance site reporting for inclusion into network studies: Observational Medical Outcomes Partnership (OMOP)^24^, Accrual to Clinical Trials (ACT)^25^, TriNetX^26^, and Patient-Centered Clinical Research Network (PCORnet).^27^

Where possible, we categorically excluded data providers rather than individual participants based on minimum data reporting requirements and robustness measures. We excluded data partners who pre-shifted dates by >45 days, reported limited death information (less than one standard deviation below mean reporting), had insufficient measurement information to calculate BMI on most of their population (>50%), and did not provide 5-digit ZIP Codes. We excluded patients with missing age and gender across all data partners.

### Data Extraction

Data were extracted on June 4, 2021, (N3C release 32) in the OMOP Common Data Model version 5.3.1.^13^ This facilitates a 2-month window for data reporting from our diagnostic cutoff (March 31, 2021) to support 30-day outcomes analyses and comprehensive reporting from data partners. All clinical concept sets were created collaboratively within the N3C Enclave, with at least one informatician and one clinical subject-matter expert reviewing each relevant concept set. Concept sets^28^ contain standardized terminology corresponding to clinical domains (e.g., LOINC, SNOMED CT, ICD-10, RxNorm). Logistic models were calculated with all C19 patients. All-cause mortality was computed on hospitalized patients as published literature suggests that the most reliable and timely death data are available in EHRs of hospitalized patients, representing 64% of all death certificates in the final quarter of 2020.^29^

### Covariates

N3C provides patient ZIP Codes for most patients (∼66% of all subjects). The majority of missing ZIP Code information is from specific data providers who elect not to provide 5-digit ZIP Codes in their data transmissions. We relied on a RUCA Codes crosswalk to match ZIP Codes and RUCA classifications based on data availability.^30^ Current RUCA codes derive from the 2010 Census and the 2006-2010 American Community Survey.^18^ We defined rural areas broadly according to the Office of Management and Budget and Federal Office of Rural Health Policy (FORHP) definitions (primary RUCA code between 1-3 urban, and 4-10 rural). Based on FORHP definitions, we divide rural areas into two categories: UAR (RUCA codes 4-5, 7-8) and NAR (RUCA codes 6, 9-10).^31^

### Outcomes

Primary outcomes are hospital admission and all-cause mortality (any reported death or a discharge to hospice) among hospitalized patients. Survival analyses assessed mortality at 30 days post-hospitalization. Secondary outcomes included length of stay, supplemental oxygen, mechanical ventilation, and major adverse cardiovascular event (MACE) or extracorporeal membrane oxygenation (ECMO).

### Statistical Analyses

Summary statistics using chi-square tests of independence were calculated on all C19 patients (Appendix A), hospitalized C19 patients (Table 1), and hospitalized non-C19 patients (Appendix B), stratified by rural-urban categories.^32^ Covariates examined include gender, age, race, ethnicity, BMI, Charlson Comorbidity Index (CCI) ^33^ composite score (a higher score indicating worse health), comorbidity categories, tobacco usage, hospitalization, and death/discharge to hospice. Mixed-effects logistic regression models were calculated for the entire C19 and hospitalized C19 groups. Both models included fixed effects for gender, race, ethnicity, BMI, age at visit start, CCI score, and rural category, with an outcome of all-cause inpatient mortality, and random effects for data partner.

**Table 1:** Baseline Characteristics Hospitalized COVID-19 Positive Population by Rurality Category, January 2020 – March 2021.

| Characteristic | Urban,<br>N = 104,051<sup>1</sup> | Urban-Adjacent<br>Rural, N = 11,121<sup>1</sup> | Nonurban-Adjacent<br>Rural, N = 2,725<sup>1</sup> | p value<sup>2</sup> |
| --- | --- | --- | --- | --- |
| **Gender** |  |  |  | <0.001 |
| Female | 53,905 (52%) | 5,597 (50%) | 1,310 (48%) |  |
| Male | 50,146 (48%) | 5,524 (50%) | 1,415 (52%) |  |
| **Age Group** |  |  |  | <0.001 |
| <18 | 4,261 (4.1%) | 540 (4.9%) | 91 (3.3%) |  |
| 18-29 | 10,199 (9.8%) | 948 (8.5%) | 194 (7.1%) |  |
| 30-49 | 24,602 (24%) | 2,117 (19%) | 400 (15%) |  |
| 50-64 | 26,583 (26%) | 2,978 (27%) | 770 (28%) |  |
| >=65 | 38,406 (37%) | 4,538 (41%) | 1,270 (47%) |  |
| **Race** |  |  |  | <0.001 |
| White | 48,067 (46%) | 7,238 (65%) | 1,974 (72%) |  |
| Black or AA | 25,858 (25%) | 2,171 (20%) | 473 (17%) |  |
| Asian or NHPI | 4,252 (4.1%) | 69 (0.6%) | <20<sup>3</sup> |  |
| Other | 772 (0.7%) | 154 (1.4%) | ~50 (1.7%)<sup>3</sup> |  |
| Missing/Unknown | 25,102 (24%) | 1,489 (13%) | 224 (8.2%) |  |
| **Ethnicity** |  |  |  | <0.001 |
| Not Hispanic or Latino | 77,099 (74%) | 9,649 (87%) | 2,487 (91%) |  |
| Hispanic or Latino | 20,611 (20%) | 1,166 (10%) | 172 (6.3%) |  |
| Missing/Unknown | 6,341 (6.1%) | 306 (2.8%) | 66 (2.4%) |  |
| **BMI Category** |  |  |  | <0.001 |
| <18.5 | 3,856 (3.7%) | 340 (3.1%) | 71 (2.6%) |  |
| 18.5-24.9 | 21,035 (20%) | 1,989 (18%) | 496 (18%) |  |
| 25-29.9 | 26,721 (26%) | 2,727 (25%) | 669 (25%) |  |
| >30 | 41,884 (40%) | 5,367 (48%) | 1,331 (49%) |  |
| Unknown/Missing | 10,555 (10%) | 698 (6.3%) | 158 (5.8%) |  |
| **Charlson Comorbidity Index Composite** |  |  |  | <0.001 |
| <1.0 | 54,937 (53%) | 5,676 (51%) | 1,328 (49%) |  |
| 1.0-2.0 | 23,002 (22%) | 2,283 (21%) | 558 (20%) |  |
| >2.0 | 26,112 (25%) | 3,162 (28%) | 839 (31%) |  |
| Composite Score (CI) | 0.00 (0.00, 3.00) | 0.00 (0.00, 3.00) | 1.00 (0.00, 3.00) |  |
| **Comorbidity Incidence** |  |  |  |  |
| Hypertension | 38,141 (37%) | 4,357 (39%) | 1,115 (41%) | <0.001 |
| Diabetes Mellitus | 23,646 (23%) | 2,814 (25%) | 720 (26%) | <0.001 |
| Myocardial Infarction | 6,337 (6.1%) | 799 (7.2%) | 210 (7.7%) | <0.001 |
| Congestive Heart Failure | 11,969 (12%) | 1,501 (13%) | 388 (14%) | <0.001 |
| Peripheral Vascular Disease | 8,964 (8.6%) | 1,166 (10%) | 284 (10%) | <0.001 |
| Stroke | 9,614 (9.2%) | 1,087 (9.8%) | 276 (10%) | 0.061 |
| Dementia | 4,395 (4.2%) | 444 (4.0%) | 129 (4.7%) | 0.2 |
| Chronic Pulmonary Disease | 17,371 (17%) | 1,933 (17%) | 482 (18%) | 0.081 |
| Rheumatologic Disease | 4,088 (3.9%) | 460 (4.1%) | 114 (4.2%) | 0.5 |
| Mild or Severe Liver Disease | 7,426 (7.1%) | 807 (7.3%) | 220 (8.1%) | 0.2 |
| Hemiplegia or paraplegia | 1,835 (1.8%) | 261 (2.3%) | <50<sup>3</sup> | <0.001 |
| Renal Disease | 13,731 (13%) | 1,716 (15%) | 476 (17%) | <0.001 |
| Any malignancy (except skin) | 10,134 (9.7%) | 1,153 (10%) | 317 (12%) | <0.001 |
| Metastatic solid tumor | 2,426 (2.3%) | 276 (2.5%) | 73 (2.7%) | 0.3 |
| HIV/AIDS | 1,039 (1.0%) | 47 (0.4%) | <20<sup>3</sup> | <0.001 |
| Multiple Comorbidities | 54,110 (52%) | 5,940 (53%) | 1,509 (55%) | <0.001 |
| Current or former smoker | 38,099 (37%) | 2,384 (21%) | 714 (26%) | <0.001 |
| **Outcomes** |  |  |  |  |
| Any Oxygen Support | 13,388 (13%) | 2,079 (19%) | 588 (22%) | <0.001 |
| Any Mechanical Ventilation | 10,087 (9.7%) | 1,867 (17%) | 477 (18%) | <0.001 |
| Hospital Readmission | 3,306 (3.2%) | 188 (1.7%) | 32 (1.2%) | <0.001 |
| ECMO or MACE | 947 (0.9%) | 174 (1.6%) | 46 (1.7%) | <0.001 |
| All-Cause Inpatient Mortality or Hospice | 11,240 (11%) | 1,763 (16%) | 455 (17%) | <0.001 |
| Time to Death in Days (CI) | 13 (6, 28) | 14 (7, 27) | 14 (5, 27) | 0.6 |
| **Quarter of Diagnosis** |  |  |  | <0.001 |
| Jan-Mar 2020 | 6,145 (5.9%) | 85 (0.8%) | 25 (0.9%) |  |
| Apr-Jun 2020 | 24,478 (24%) | 1,507 (14%) | 438 (16%) |  |
| Jul-Sep 2020 | 15,851 (15%) | 2,114 (19%) | 482 (18%) |  |
| Oct-Dec 2020 | 32,856 (32%) | 4,337 (39%) | 995 (37%) |  |
| Jan-Mar 2021 | 24,721 (24%) | 3,078 (28%) | 785 (29%) |  |

<sup>1</sup> Statistics presented: n (%)
<sup>2</sup> Statistical tests performed: chi-square test of independence
<sup>3</sup> Censored to remove small cell count or potential reidentification of small cell count

We assessed model specification using a methodology that checked for specification change influence on the estimate rural versus urban effect. Among the possible specification changes assessed were (a) transformations of main effects included in the model, (b) all two-way interactions of main effects included in the model, (c) potential addition of available comorbidities not included in the model, and (d) include of data providing organization as a fixed or random effect. In addition, a stepwise assessment approach was used to determine if combinations of any such potential changes resulted in change in the estimated rural versus urban effect. The only specification change identified was the need to include the data provider organization as a fixed or random effect, with the choice of effect type (fixed versus random) found to be irrelevant. As a result, the initial model was modified to include data providing organization as a random effect. This helps mitigate the possibility that our final model’s estimated rural effects are artifacts of mortality differences across those organizations’ patient populations.

The risk adjustment process employed in our modeling used information about pre-COVID comorbidities. As patient data in N3C differ in availability of pre-COVID clinical data, ranging from none to two years of pre-COVID clinical data, we examined the possibility that estimated rural effects stemmed from rural and urban patients differing in extent of pre-COVID comorbidity information.

The variables associated with worse outcomes in the logistic model – rurality, Charlson Comorbidity Index, age, and period of SARS-CoV-2 diagnosis – were secondarily evaluated using Kaplan-Meier estimates of the overall time to death, starting from hospital admission, censored at 30 days.

## Results

### Demographics

Our final cohort included data from 27 data partners (Figure 1) with 573,018 C19 diagnosed,117,897 hospitalized C19, and 450,725 hospitalized non-C19 patients. Patient demographics in the entire C19 cohort demonstrated rural inhabitants to be older and less racially and ethnically diverse in all groups examined, i.e., the hospitalized C19 positive group (Table 1), non-hospitalized Covid19 positive group (Appendix A), and hospitalized Covid19 negative comparison group (Appendix B).

**Figure 1:**
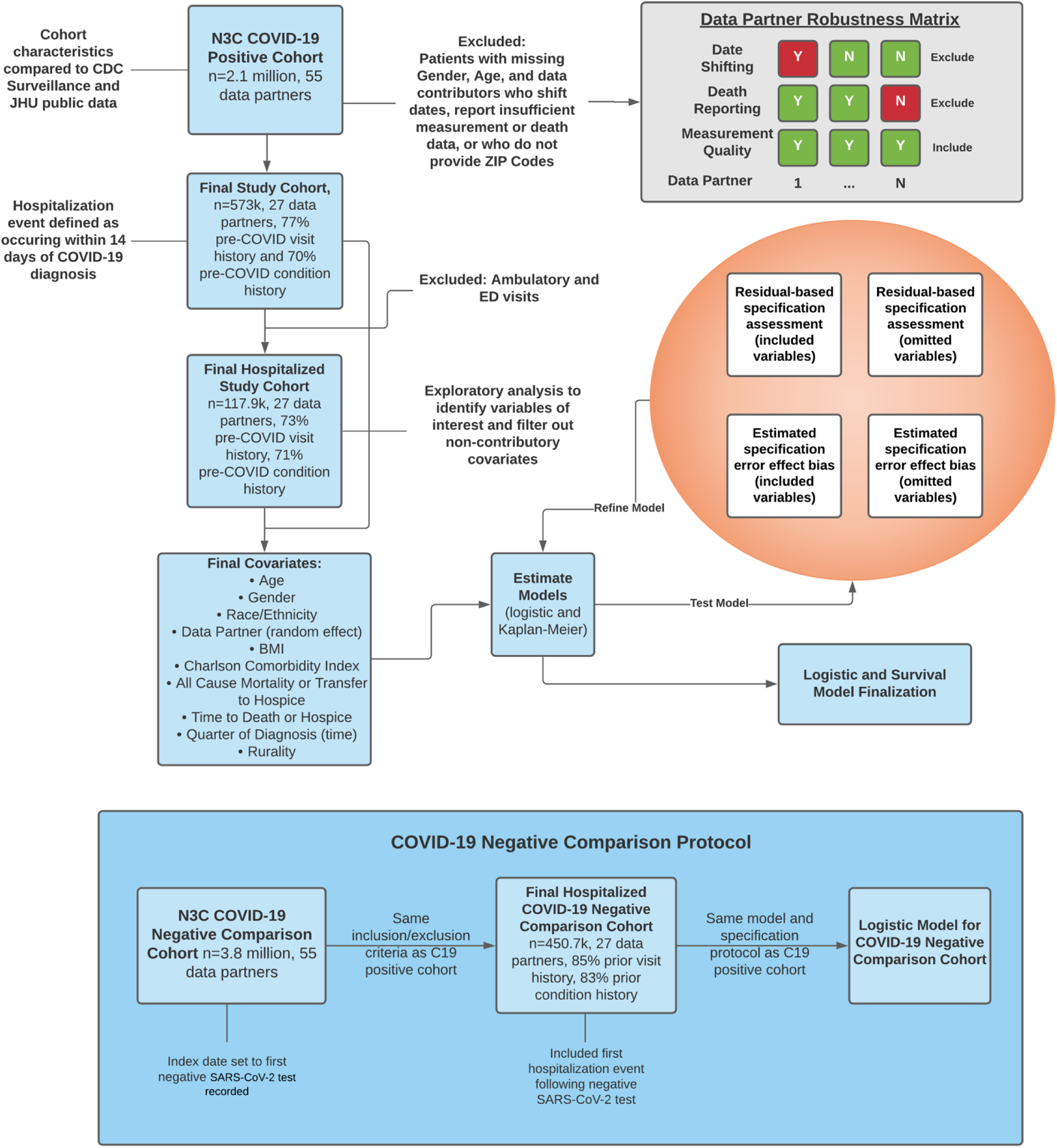
Data Analysis Plan. Figure 1 documents the data analysis plan, including steps for inclusion and exclusion of data partners based on availability of ZIP Codes, whether they pre-shifted dates by over 45 days, and robustness based on covariates of interest (measurement domain to calculate BMI and death domain for primary outcome). Our Data Robustness Matrix includes determining fact counts in each domain of interest (death, measurement) and excluding sites who were one standard deviation below the mean reporting percentage in our initial cohort.

### Underlying Health Disparities and Observable Vulnerabilities

Among all patients, rural populations had higher rates of comorbidities across 14 of the 15 comorbidity categories (Appendix C) and had notably higher rates of obesity (Table.1). We also noted that among our C19 cohorts, between 65% and 80% of patients in each category had prior visit history and between 63% and 74% had prior conditions reported in the pre-COVID period (Figure 1), suggesting that our data robustness matrix sufficiently captured patients with high-fidelity data.

Assessing date of hospitalization, as a proxy for changes in clinical practice and treatment, we found that rural C19 patients were more likely to be diagnosed, and subsequently hospitalized later in the pandemic (Appendix C) when treatment practices were leading to better outcomes. Urban dwellers had higher caseloads in quarter 2 (17% urban, 10% UAR, 10% NAR; p<0.001) and quarter 3 (18% urban, 16% UAR, 15% NAR; p<0.001) of 2020. Urban patients also had higher hospitalization rates in quarter 2 (24% urban, 14% UAR, 16% NAR; p<0.001), but more rural patients were hospitalized in quarter 3 (15% urban, 19% UAR, 18% NAR; p<0.001).

### Hospitalization

Persons in rural areas were more likely to be hospitalized, UAR (OR 1.62, 95% CI, 1.58, 1.66) and NAR (OR 1.61, 95% CI, 1.54-1.69), compared to urban dwellers (Figure 3.A). After adjusting for differences in gender, race, ethnicity, BMI, age, CCI, and quarter of diagnosis, C19 patients in rural areas had approximately a 40% increased risk of hospitalization, UAR (AOR 1.41, 95% CI, 1.37-1.45) and NAR (AOR 1.42, 95% CI, 1.35-1.50) (Figure 3.B).

**Figure 2:**
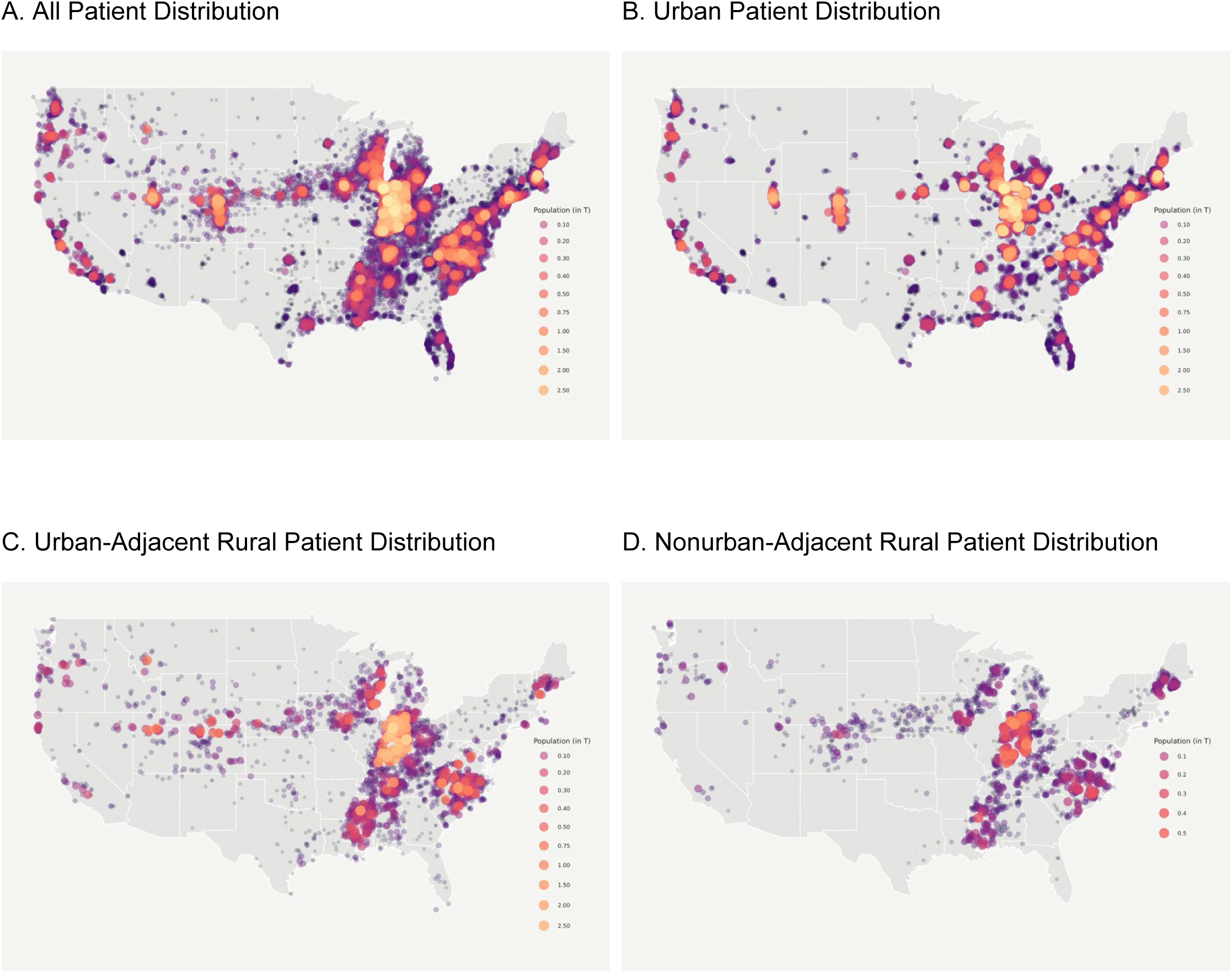
N3C Patient Distribution. Figure 2 shows the geospatial distribution of the N3C COVID-19 positive population. N3C contains data from 55 data contributors from across the United States, 40 of whom include sufficient location information to spatially map by ZIP Code centroid. Of those sites, we selected 27 whose data met our minimum robustness qualifications for inclusion in our study. This bubble map is to scale with larger bubbles representing more patients. Figure 2.A. shows all N3C patients. Figure 2.B. shows only urban N3C distribution. Figure 2.C. shows urban-adjacent rural patient distribution. Figure 2.D. shows nonurban-adjacent rural patient distribution, which represents the most isolated patients in N3C.

**Figure 3:**
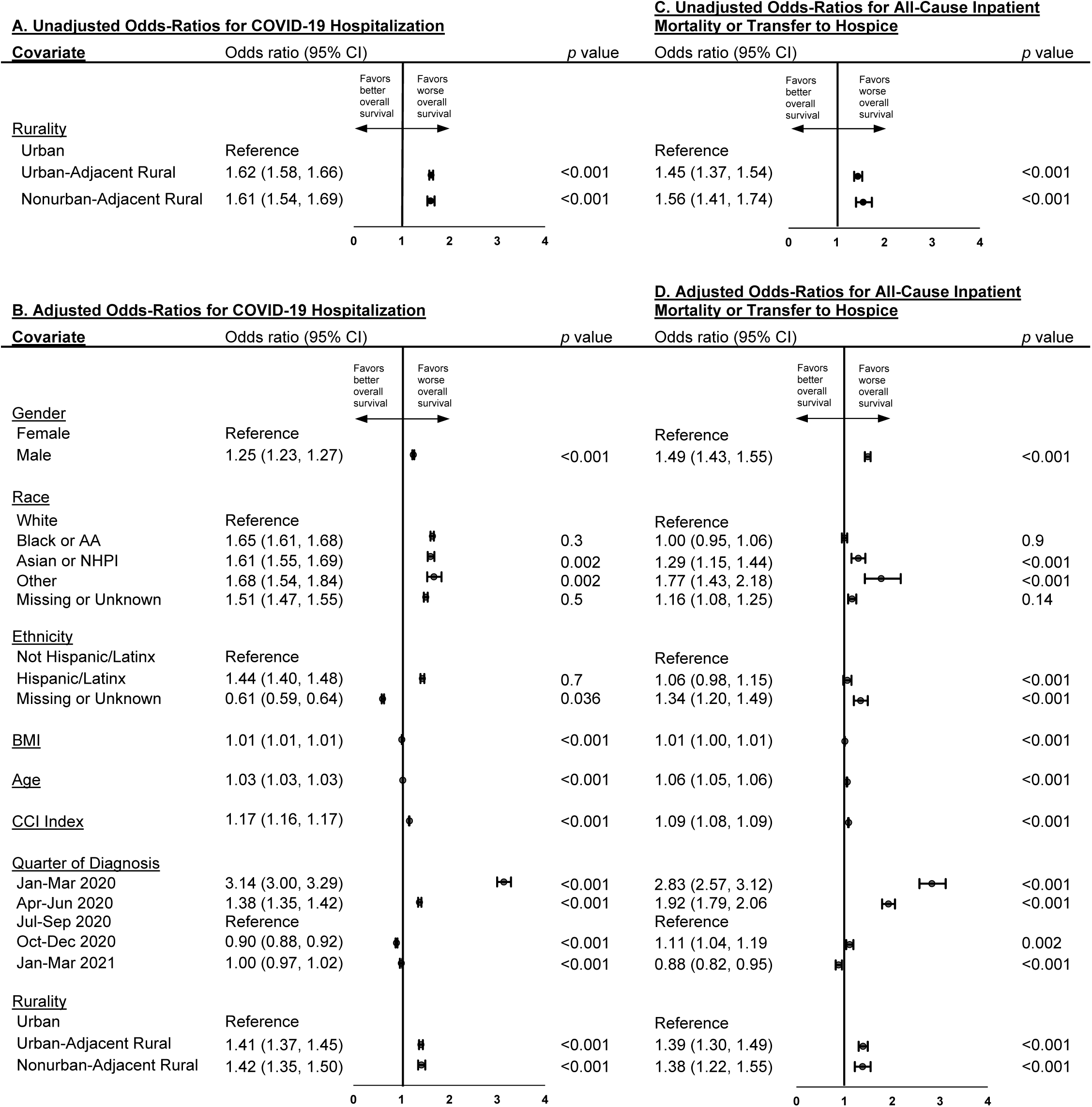
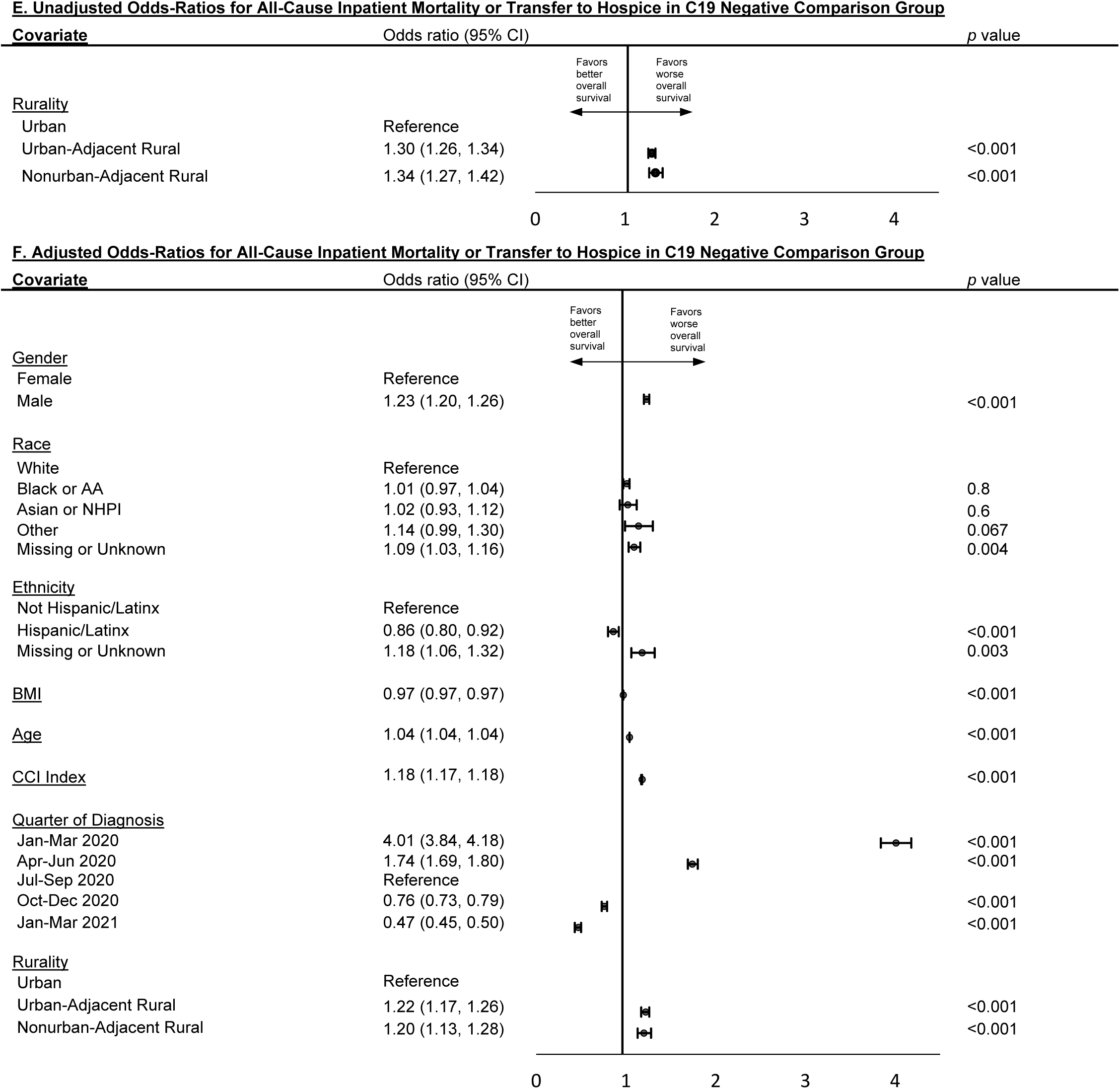
Forest Plot Showing the Unadjusted and Adjusted Odds Ratios for Hospitalization and All-Cause Inpatient Mortality or Transfer to Hospice by Rural Category in C19 Positive Cohort, January 2020 – March 2021. Figure 3 shows the unadjusted odds and adjusted ratios for being hospitalized (A and B) and dying or being transferred to hospice after hospitalization for the COVID-19 infected population (C and D) and uninfected population (E and F) in N3C. Risk is similar between adjusted and unadjusted models, suggesting a real impact of rurality on hospitalization and all-cause mortality. Adjusted models include adjustments for gender, race, ethnicity, BMI, age, Charlson Comorbidity Index (CCI) composite score, rurality, and quarter of diagnosis. Data provider is included as a random effect in all models to account for differences across source data systems.

### Mortality

The model estimates of all-cause inpatient mortality or transfer to hospice after C19 diagnosis were significantly greater for rural compared to urban patients, (OR 1.45, 95% CI, 1.37, 1.54) and NAR (OR 1.56, 95% CI, 1.41-1.74) (Figure 3.C). After adjusting for differences in gender, race, ethnicity, BMI, age, CCI, and quarter of diagnosis, mortality remained approximately 40% greater for rural C19 hospitalized patients, UAR (AOR 1.39, 95% CI, 1.30-1.49) and NAR (AOR 1.38, 95% CI, 1.22-1.55) (Figure 3.D).

Kaplan Meier survival curves demonstrate significantly higher mortality 30 days after hospitalization among rural C19 patients compared to their urban counterparts. (Figure 4.A). Hospitalized C19 patients with more comorbidities, older age, and diagnosis earlier in the pandemic demonstrated significantly higher mortality (Figure 4.B, 4.C, 4.D).

**Figure 4:**
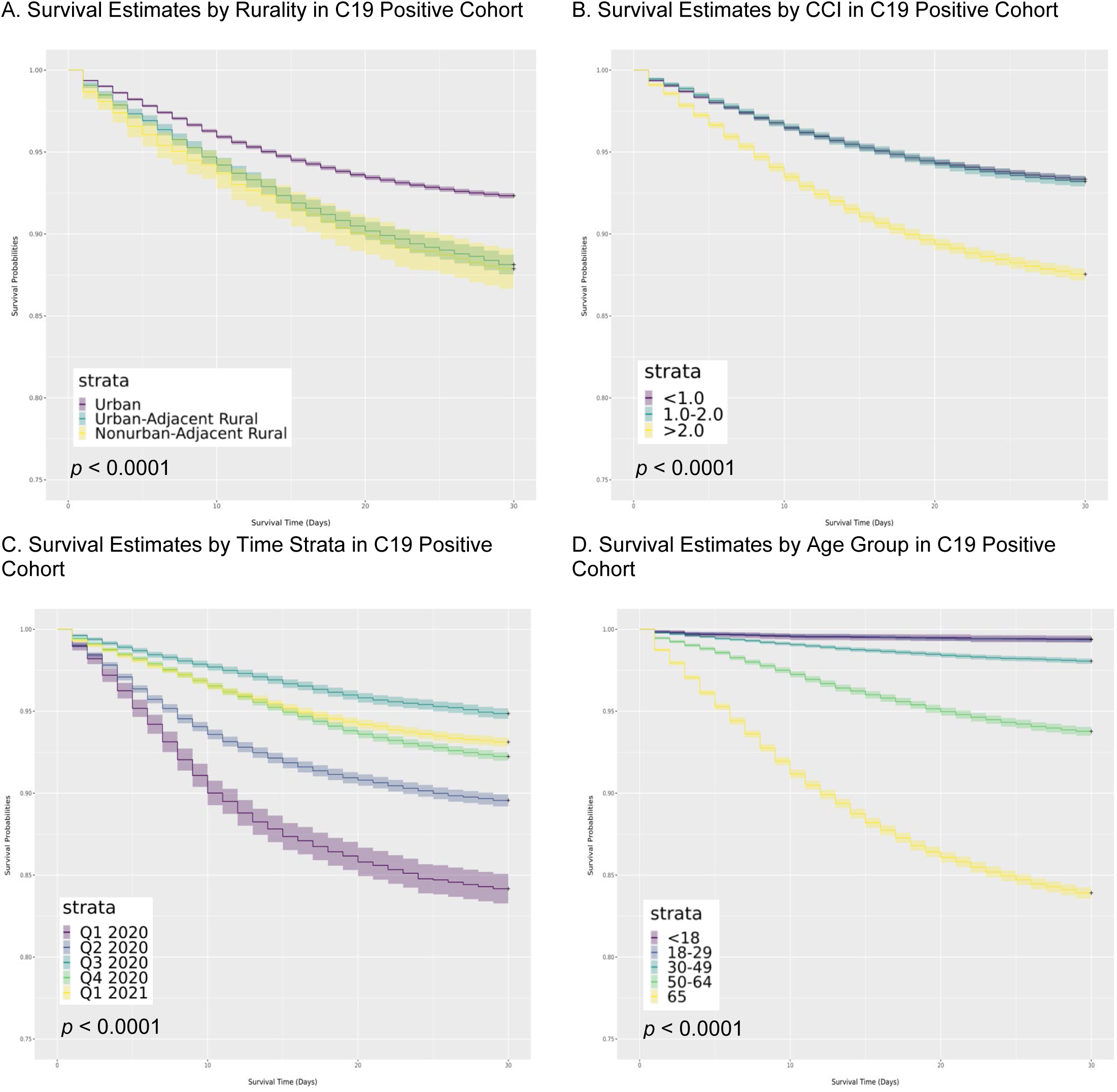
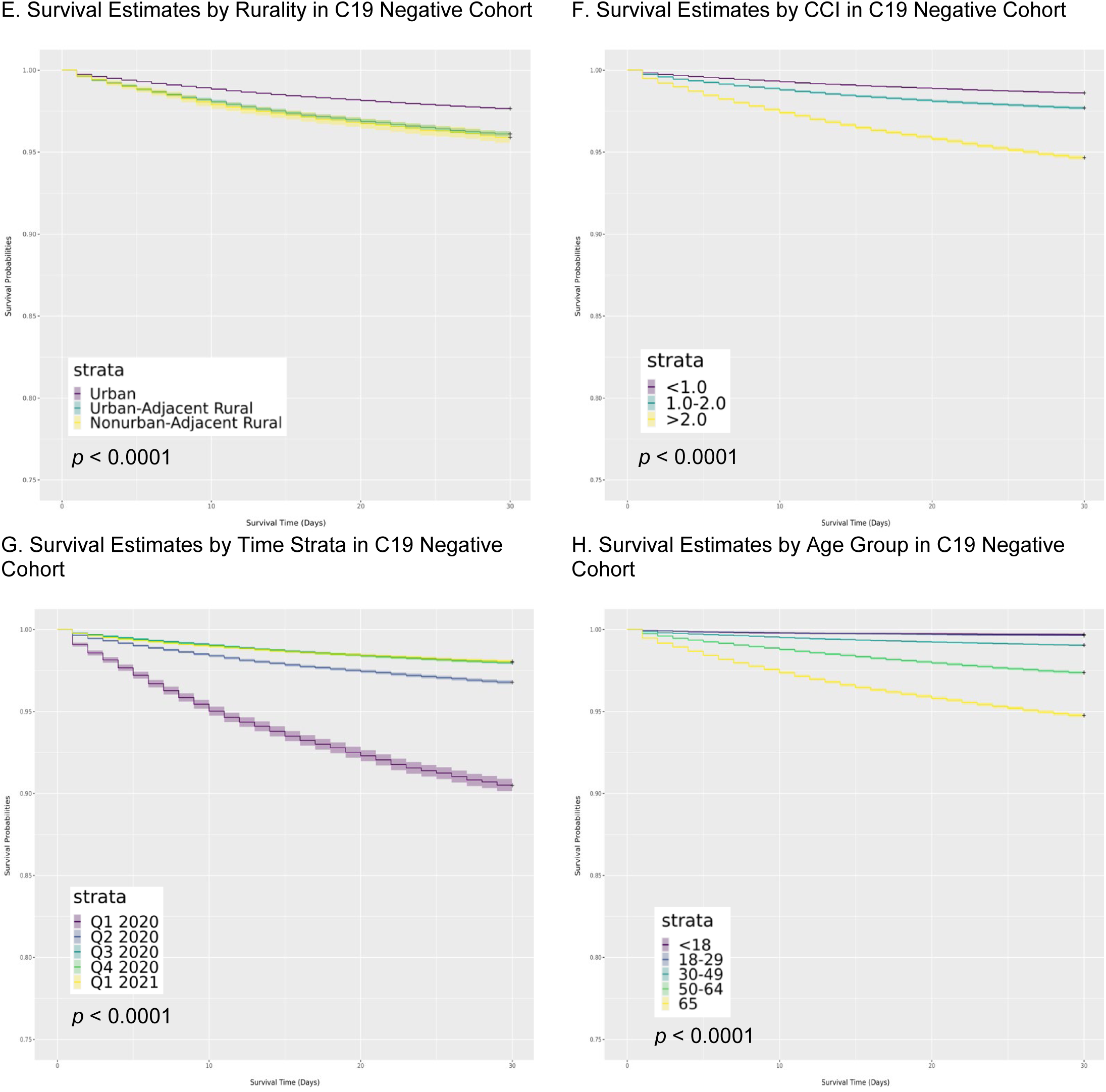
Kaplan-Meier Survival Curves in Hospitalized Patients Over 30 Days from Day of Admission. Appendix A shows Kaplan-Meier survival estimates in both COVID-19 infected and uninfected hospitalized cohorts in N3C by rural category (A and E, respectively), Charlson Comorbidity Index (B and F, respectively), Quarter of Diagnosis or earliest negative SARS-CoV-2 lab test (C and G, respectively), and Age Group (D and H, respectively) from hospital admission through day 30. Events were censored at day 30 based on incidence of death or transfer to hospice care. These four factors had the highest predictive power of the covariates evaluated in this study.

### SARS-CoV-2 Uninfected Comparison Cohort

The same analyses on the hospitalized, uninfected comparison group demonstrated similar, albeit attenuated, results as the C19 positive group. This cohort had similar background characteristics (Appendix B) and similar likelihood of being hospitalized throughout the study period. Hospitalized uninfected patients in rural areas had significantly greater mortality compared with urban uninfected patients, UAR (OR 1.30, 95% CI, 1.26-1.34) and NAR (OR 1.34, 95% CI, 1.27-1.42) compared to urban dwellers (Figure 3.E). After adjusting for differences in gender, race, ethnicity, BMI, age, CCI, and quarter of diagnosis, rurality was still associated with approximately 20% increased odds of mortality in UAR (AOR 1.22, 95% CI, 1.17-1.26) and NAR (AOR 1.20, 95% CI, 1.13-1.28) (Figure 3.F).

Kaplan Meier survival curves show similar differences across the SARS-CoV-2 uninfected cohort as the infected cohort, albeit with a 3-fold higher likelihood of survival in all categories. Uninfected rural patients face significantly higher mortality 30 days after hospitalization compared to their urban counterparts (Figure 4.E). As in the infected cohort, uninfected hospitalized persons with more comorbidities, older age, and hospitalization earlier in the pandemic demonstrated higher mortality (Figure 4.F, 4.G, 4.H).

### Secondary Outcomes

Treatment with ECMO or occurrence of MACE were relatively infrequent among C19 patients but ranged from 0.9% in urban to 1.6% in UAR and 1.7% in NAR (p<0.001). Invasive mechanical ventilation and oxygen support were also significantly more frequent among rural than among urban patients (Table 1). However, the mean time to death was similar ranging between 12-14 days after C19 diagnosis among those hospitalized. Urban patients were more likely to be readmitted to the hospital at any point after initial hospitalization in both the infected and uninfected cohorts.

## Discussion

In this retrospective cohort study from a large representative data base of both C19 positive and negative patients, we found significantly higher mortality rates among hospitalized rural C19 patients. Mortality was approximately 40% higher among rural C19 patients after adjustment for age, gender, race, ethnicity, BMI, Charlson Comorbidity Index composite score, date of diagnosis, and differences derived from the data contributor.

The SARS-CoV-2 pandemic provides an important lens to examine all-cause mortality by area of residence. SARS-CoV-2 is a new virus, rapidly transmitted in an immunologically naïve human population and initially without known effective treatment. The rapidity with which first urban then rural communities in the US experienced surges of SARS-CoV-2, led us to hypothesize that an urban-rural mortality differential would not be observed. That, however, is not the case. Although rural populations are older and have higher rates of comorbidities,^34^ including diabetes mellitus and obesity^35^ which have been associated with increased disease severity and death in SARS-CoV-2 infection, adjustment for these factors did not change the finding of higher rural mortality. Notably, mortality among hospitalized patients without C19 diagnoses, after adjustment for several factors, was also significantly higher in rural compared to urban areas, a finding consistent with previous studies.^36, 37^

The gradient of risk for chronic diseases and mortality between urban and rural inhabitants is a relatively recent development in the United States. Prior to 1980, mortality rates in rural and urban areas of the US were comparable. Since then, mortality rates in rural America have exceeded urban rates, and the gap has accelerated since 1999, even when adjusted for age.^38,34,35^ This mortality disparity has been referred to as “the nonmetropolitan penalty,” ^39,^ and some experts believe that structural urbanism is widening the gap.^40^ Similar findings are noted among nonmetropolitan counties for common causes of death, including heart disease, cancer, chronic lower respiratory disease, unintentional injury, and stroke.^6^

Given the modeling objectives, it would be illogical to adjust the rural versus urban comparison for clinical severity, such as clinical severity at the time of hospital admission. If a rural versus urban effect exists, it almost certainly would lead to rural versus urban differences in clinical severity at admission. Therefore, adjusting for clinical severity at admission would result in adjusting away the effect which the modeling seeks to evaluate. Our modeling showed that the rural versus urban effect was larger among patients hospitalized for COVID than among patients hospitalized for non-COVID reasons. This suggests that rurality played a greater relative role in COVID outcomes than it plays in the outcomes resulting from most other (non-COVID) hospitalizations.

The etiology of this “penalty” is likely multi-factorial. Poverty, unemployment, and lower levels of educational attainment are more prevalent in rural areas, and these factors may partially explain the urban-rural mortality disparity. Diminished access to care has also been described as contributing to rural-urban mortality differences. One study concluded that having one or more specialist visits during the previous year was associated with 16.6% lower mortality for those with chronic conditions.^39,41^ Among counties with defined shortages in primary care delivery, 56% are nonmetropolitan while 19% are metropolitan, according to data reported by the Health Resources and Services Administration.^42^

Although limited research has been done on socioeconomic risk factors and case-fatality rates over the first year of the SARS-Cov-2 pandemic, a recent study from Japan observed higher C19-related mortality in prefectures with the lowest household incomes.^43^ A recent study conducted using spatial models in the US found rurality to be one of several ecologic determinants of C19 mortality.^44^ Our results again raise the question of why rural populations experience higher mortality rates after adjustment for multiple factors, even with a new pathogen such as SARS-CoV-2. To what extent delays in care contribute to increased SARS-CoV-2 related mortality among rural populations is unclear as is the potential impact of environmental risk factors in rural areas. The increase in hospitalization and hospitalized mortality in our uninfected cohort demonstrates systemic discrepancies in rural-urban outcomes in tertiary care centers that are likely exacerbated by C19.

Identifying and understanding potential causes of urban-rural health disparities including mortality across both chronic and acute conditions may inform study of rational and cost-effective mitigation strategies. Others^45, 46, 47^ have demonstrated that attribution of rurality does not provide a one-size-fits-all means to prescribe approaches to reduce health disparities. There are many other possible contributors to the observed disparity, highlighting the urgent need for a robust research agenda that will address the root causes which may differ by geographic region.

While confirming worse baseline health status and outcomes of rural dwellers from chronic conditions, the N3C data also suggest rural dwellers have incremental vulnerability to C19 as an acute health condition. The disparately adverse C19 acute outcomes among rural dwellers do not appear to be fully explained by the worse baseline health measures. The C19 pandemic appears to both highlight and extend the apparent relative vulnerability of rural dwellers to acute health conditions. Recent scholarship points to a dynamic, ecological model for addressing rural adversity based on tailored approaches to individual community needs.^48^ Additional observational and experimental research is needed to potentially identify practical evidence-based steps to improve health outcomes among rural dwellers for both acute and chronic health conditions.

### Limitations

N3C is an observational registry compiling data from multiple diverse participating sites. Therefore, some information may be entirely or partially and non-randomly missing from the database in rural versus urban residents. In our C19 positive cohort, we report incidence of comorbidities in more than two-thirds of our study population, which is similar to those reported in a COVID-19 study across OCHIN, a network of 396 community health centers across 14 states.^49^ Nonetheless, we examined the possibility that estimated rural effects stemmed from rural and urban patients differing in extent of pre-COVID comorbidity information and found this not to be the case. To some degree, such potential bias can be partly assessed by future analyses, which would include data based on all diagnosed C19+ populations in geographic areas, rather than data limited to C19 patients who received care at N3C collaborating provider systems.^50^ A second potential confounder is that rural residents with C19 may have been diagnosed later in their disease course than urban residents, leading to a greater risk of serious complications. Additionally, healthcare organizations contributing data to N3C may have cared for more severely ill patients, providing a potential source of bias.

Other limitations of the N3C data source include data aggregated from different health systems with different local practices, regulations, and data models, resulting in potential reporting differences, despite our application of data robustness checks. Additionally, the comparison group is limited only to those pre-matched at the site level from patients who have had a confirmed negative SARS-CoV-2 test. Although 54% of American Indian and Alaska Native populations in the US live in rural areas and have been disproportionately affected by Covid19,^51, 52^ their racial demographic is unavailable for explicit study in accordance with tribal sovereignty policies. Finally, these analyses are limited to residents in the US and may not be generalizable to other countries.

### Conclusions

Hospitalization and death were significantly more frequent among rural C19 patients than their urban counterparts after adjusting for multiple factors, including age, sex, race, and comorbidities. An analysis of adjusted mortality among hospitalized SARS-CoV-2 uninfected patients demonstrated similar results. These data provide evidence-based documentation of rural health disparities. Further research is needed to understand this disparity for both acute and chronic health conditions.

## Data Availability

All data included in this manuscript are available publicly in the National COVID Cohort Collaborative (N3C). Data utilized in this study are from release 32. All code and concepts described herein are available on our project GitHub repository: https://github.com/National-COVID-Cohort-Collaborative/CS-Rural-Health/tree/main/rural-mortality-and-hospitalization

https://covid.cd2h.org/

## Appendix A: Baseline Characteristics of All N3C COVID-19 Positive Population by Rurality Category, January 2020 – March 2021

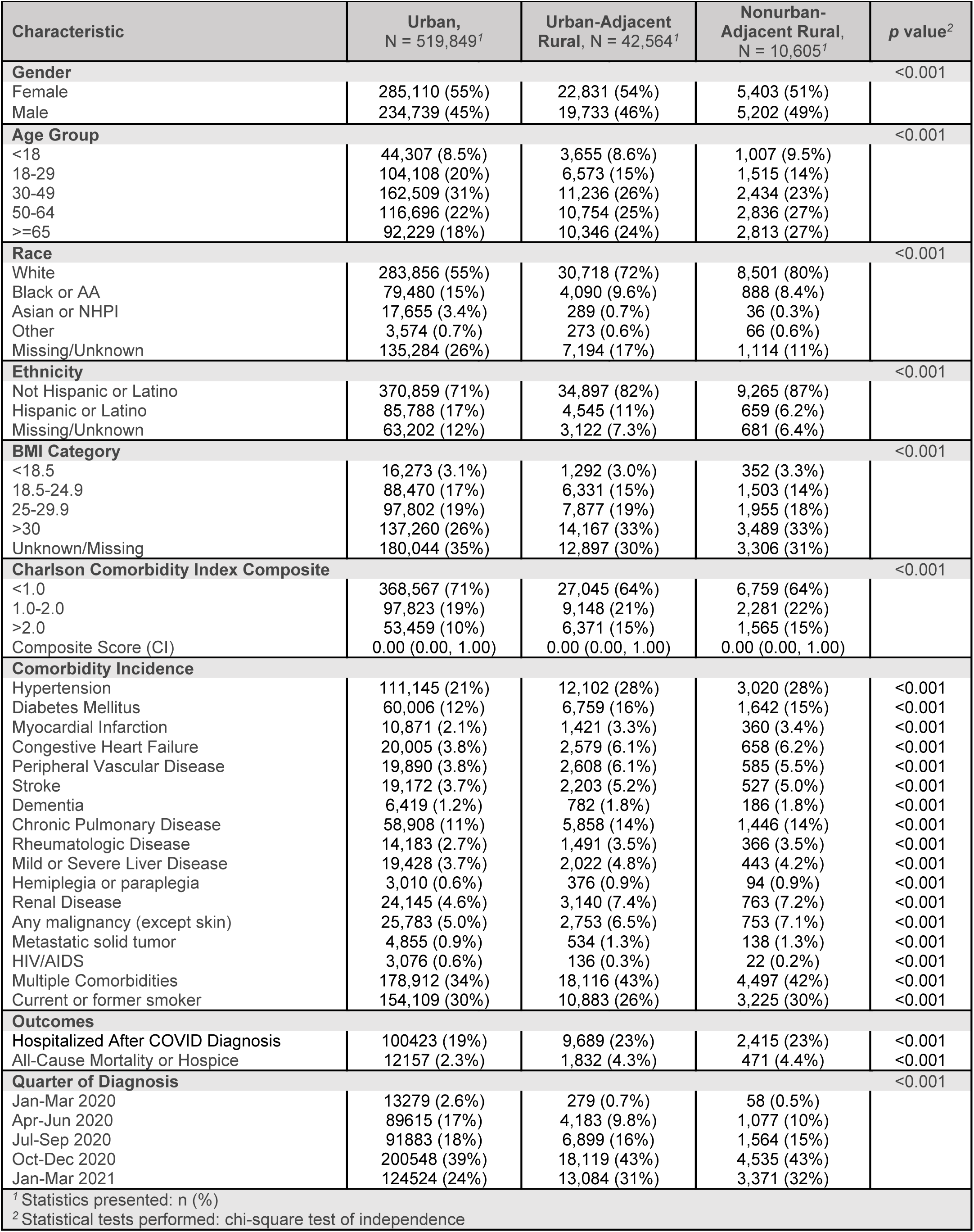

| Characteristic | Urban,<br>N = 519,849<sup>1</sup> | Urban-Adjacent<br>Rural, N = 42,564<sup>1</sup> | Nonurban-Adjacent Rural,<br>N = 10,605<sup>1</sup> | p value<sup>2</sup> |
| --- | --- | --- | --- | --- |
| **Gender** |  |  |  | <0.001 |
| Female | 285,110 (55%) | 22,831 (54%) | 5,403 (51%) |  |
| Male | 234,739 (45%) | 19,733 (46%) | 5,202 (49%) |  |
| **Age Group** |  |  |  | <0.001 |
| <18 | 44,307 (8.5%) | 3,655 (8.6%) | 1,007 (9.5%) |  |
| 18-29 | 104,108 (20%) | 6,573 (15%) | 1,515 (14%) |  |
| 30-49 | 162,509 (31%) | 11,236 (26%) | 2,434 (23%) |  |
| 50-64 | 116,696 (22%) | 10,754 (25%) | 2,836 (27%) |  |
| >=65 | 92,229 (18%) | 10,346 (24%) | 2,813 (27%) |  |
| **Race** |  |  |  | <0.001 |
| White | 283,856 (55%) | 30,718 (72%) | 8,501 (80%) |  |
| Black or AA | 79,480 (15%) | 4,090 (9.6%) | 888 (8.4%) |  |
| Asian or NHPI | 17,655 (3.4%) | 289 (0.7%) | 36 (0.3%) |  |
| Other | 3,574 (0.7%) | 273 (0.6%) | 66 (0.6%) |  |
| Missing/Unknown | 135,284 (26%) | 7,194 (17%) | 1,114 (11%) |  |
| **Ethnicity** |  |  |  | <0.001 |
| Not Hispanic or Latino | 370,859 (71%) | 34,897 (82%) | 9,265 (87%) |  |
| Hispanic or Latino | 85,788 (17%) | 4,545 (11%) | 659 (6.2%) |  |
| Missing/Unknown | 63,202 (12%) | 3,122 (7.3%) | 681 (6.4%) |  |
| **BMI Category** |  |  |  | <0.001 |
| <18.5 | 16,273 (3.1%) | 1,292 (3.0%) | 352 (3.3%) |  |
| 18.5-24.9 | 88,470 (17%) | 6,331 (15%) | 1,503 (14%) |  |
| 25-29.9 | 97,802 (19%) | 7,877 (19%) | 1,955 (18%) |  |
| >30 | 137,260 (26%) | 14,167 (33%) | 3,489 (33%) |  |
| Unknown/Missing | 180,044 (35%) | 12,897 (30%) | 3,306 (31%) |  |
| **Charlson Comorbidity Index Composite** |  |  |  | <0.001 |
| <1.0 | 368,567 (71%) | 27,045 (64%) | 6,759 (64%) |  |
| 1.0-2.0 | 97,823 (19%) | 9,148 (21%) | 2,281 (22%) |  |
| >2.0 | 53,459 (10%) | 6,371 (15%) | 1,565 (15%) |  |
| Composite Score (CI) | 0.00 (0.00, 1.00) | 0.00 (0.00, 1.00) | 0.00 (0.00, 1.00) |  |
| **Comorbidity Incidence** |  |  |  |  |
| Hypertension | 111,145 (21%) | 12,102 (28%) | 3,020 (28%) | <0.001 |
| Diabetes Mellitus | 60,006 (12%) | 6,759 (16%) | 1,642 (15%) | <0.001 |
| Myocardial Infarction | 10,871 (2.1%) | 1,421 (3.3%) | 360 (3.4%) | <0.001 |
| Congestive Heart Failure | 20,005 (3.8%) | 2,579 (6.1%) | 658 (6.2%) | <0.001 |
| Peripheral Vascular Disease | 19,890 (3.8%) | 2,608 (6.1%) | 585 (5.5%) | <0.001 |
| Stroke | 19,172 (3.7%) | 2,203 (5.2%) | 527 (5.0%) | <0.001 |
| Dementia | 6,419 (1.2%) | 782 (1.8%) | 186 (1.8%) | <0.001 |
| Chronic Pulmonary Disease | 58,908 (11%) | 5,858 (14%) | 1,446 (14%) | <0.001 |
| Rheumatologic Disease | 14,183 (2.7%) | 1,491 (3.5%) | 366 (3.5%) | <0.001 |
| Mild or Severe Liver Disease | 19,428 (3.7%) | 2,022 (4.8%) | 443 (4.2%) | <0.001 |
| Hemiplegia or paraplegia | 3,010 (0.6%) | 376 (0.9%) | 94 (0.9%) | <0.001 |
| Renal Disease | 24,145 (4.6%) | 3,140 (7.4%) | 763 (7.2%) | <0.001 |
| Any malignancy (except skin) | 25,783 (5.0%) | 2,753 (6.5%) | 753 (7.1%) | <0.001 |
| Metastatic solid tumor | 4,855 (0.9%) | 534 (1.3%) | 138 (1.3%) | <0.001 |
| HIV/AIDS | 3,076 (0.6%) | 136 (0.3%) | 22 (0.2%) | <0.001 |
| Multiple Comorbidities | 178,912 (34%) | 18,116 (43%) | 4,497 (42%) | <0.001 |
| Current or former smoker | 154,109 (30%) | 10,883 (26%) | 3,225 (30%) | <0.001 |
| **Outcomes** |  |  |  |  |
| Hospitalized After COVID Diagnosis | 100,423 (19%) | 9,689 (23%) | 2,415 (23%) | <0.001 |
| All-Cause Mortality or Hospice | 12,157 (2.3%) | 1,832 (4.3%) | 471 (4.4%) | <0.001 |
| **Quarter of Diagnosis** |  |  |  | <0.001 |
| Jan-Mar 2020 | 13,279 (2.6%) | 279 (0.7%) | 58 (0.5%) |  |
| Apr-Jun 2020 | 89,615 (17%) | 4,183 (9.8%) | 1,077 (10%) |  |
| Jul-Sep 2020 | 91,883 (18%) | 6,899 (16%) | 1,564 (15%) |  |
| Oct-Dec 2020 | 200,548 (39%) | 18,119 (43%) | 4,535 (43%) |  |
| Jan-Mar 2021 | 124,524 (24%) | 13,084 (31%) | 3,371 (32%) |  |

<sup>1</sup> Statistics presented: n (%)
<sup>2</sup> Statistical tests performed: chi-square test of independence

## Appendix B: Baseline Characteristics of All Hospitalized COVID-19 Negative Patients by Rurality Category, January 2020 – March 2021

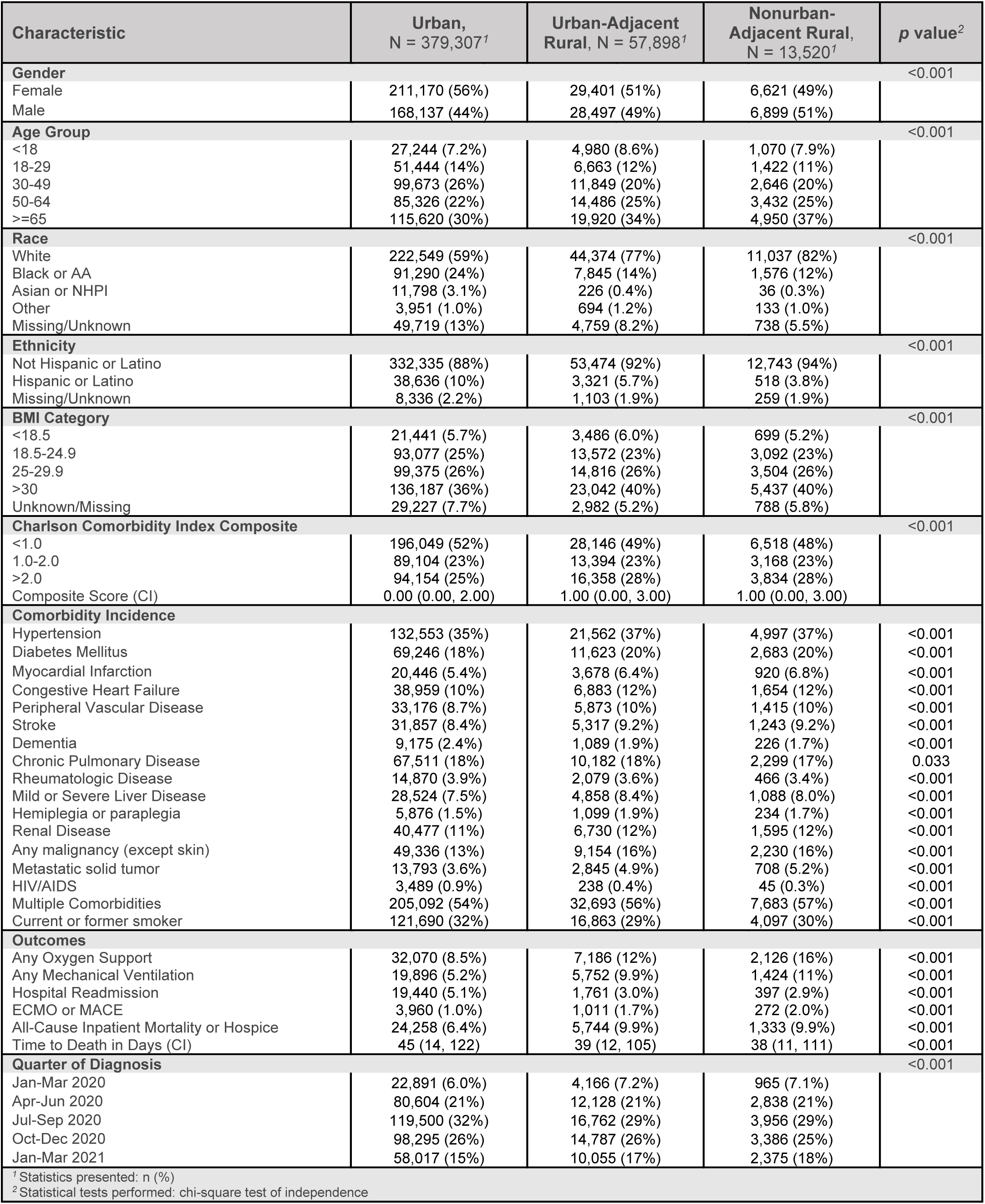

## Appendix C: Quarter of Diagnosis, COVID-19 therapies, BMI Categories, Age Group, and Comorbidities by Rural Category

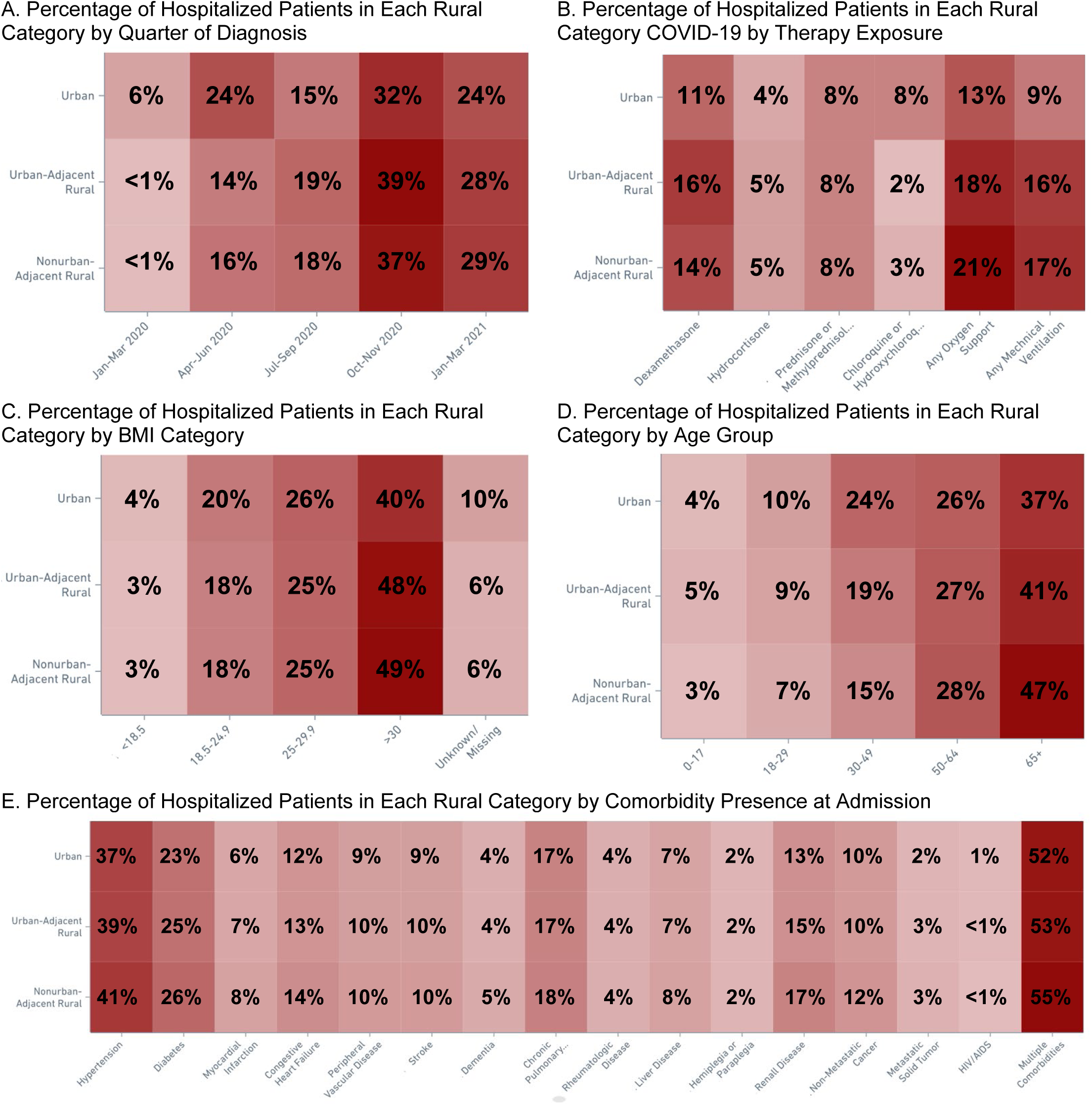
Appendix C shows differences in key areas by rurality category, including quarter of diagnosis (A) therapy exposure (B), BMI category (C) age group (D) and comorbidity incidence (E). Overall, common risk factors associated with poorer outcomes are present in the urban-adjacent and nonurban-adjacent rural cohorts (A, C, D, E), but the same cohorts receive similar treatment (B).

## Appendix D: US Summary Table of COVID-19 Caseload and Case Fatality Compared with N3C Using Public Datasets, January 2020 – March 2021

N3C Summary by RUCA Categories
| Rural Urban Commuting Area (RUCA) Categories |  | Cases<sup>1</sup> | Deaths<sup>1</sup> | Case Fatality<sup>1</sup> | Caseload<sup>1</sup> |
| --- | --- | --- | --- | --- | --- |
| Urban | RUCA 1: Metropolitan area core: primary flow within an urbanized area (UA) | 969,703 | 17,472 | 1.80% | 74.53% |
|  | RUCA 2: Metropolitan area high commuting: primary flow 30% or more to a UA | 98,361 | 1,811 | 1.84% | 7.56% |
|  | RUCA 3: Metropolitan area low commuting: primary flow 10% to 30% to a UA | 9,183 | 151 | 1.64% | 0.71% |
| Urban-Adjacent Rural | RUCA 4: Micropolitan area core: primary flow within an Urban Cluster of 10,000 to 49,999 (large UC) | 107,855 | 1,933 | 1.79% | 8.29% |
|  | RUCA 5: Micropolitan high commuting: primary flow 30% or more to a large UC | 22,021 | 446 | 2.03% | 1.69% |
|  | RUCA 7: Small town core: primary flow within an Urban Cluster of 2,500 to 9,999 (small UC) | 7,179 | 143 | 1.99% | 0.55% |
|  | RUCA 8: Small town high commuting: primary flow 30% or more to a small UC | 48,900 | 1,168 | 2.39% | 3.76% |
| Nonurban-Adjacent Rural | RUCA 6: Micropolitan low commuting: primary flow 10% to 30% to a large UC | 3,941 | 171 | 4.34% | 0.30% |
|  | RUCA 9: Small town low commuting: primary flow 10% to 30% to a small UC | 5,489 | 85 | 1.55% | 0.42% |
|  | RUCA 10: Rural areas: primary flow to a tract outside a UA or UC | 28,467 | 670 | 2.35% | 2.19% |
| **Totals** |  | **1,301,099** | **24,050** | **1.85%** | **100.00%** |
| 1. Represents the 36 N3C sites that contribute ZIP Code data at the person level that can be mapped to RUCA Codes and the removes 4 additional sites who do not report reliable death data (defined as including death data for >=0.25% of patients with COVID-19). |  |  |  |  |  |

Johns Hopkins University Summary by RUCC Categories
| Rural-Urban Continuum Code (RUCC) Categories |  | Cases<sup>1</sup> | Deaths<sup>1</sup> | Case Fatality<sup>1</sup> | Caseload<sup>1</sup> | Vaccination Percentage<sup>2</sup> |
| --- | --- | --- | --- | --- | --- | --- |
| Urban | Counties in metro areas of 1 million population or more | 16,809,750 | 303,159 | 1.80% | 55.77% | 15.13% |
|  | Counties in metro areas of 250,000 to 1 million population | 6,297,868 | 107,373 | 1.70% | 20.90% | 15.66% |
|  | Counties in metro areas of fewer than 250,000 population | 2,745,009 | 48,773 | 1.78% | 9.11% | 15.47% |
| Urban-Adjacent Rural | Urban population of 20,000 or more, adjacent to a metro area | 1,234,917 | 23,384 | 1.89% | 4.10% | 15.09% |
|  | Urban population of 2,500 to 19,999, adjacent to a metro area | 471,619 | 8,585 | 1.82% | 1.56% | 15.98% |
|  | Completely rural or less than 2,500 urban population, adjacent to a metro area | 1,396,410 | 29,550 | 2.12% | 4.63% | 14.50% |
| Nonurban-Adjacent Rural | Urban population of 20,000 or more, not adjacent to a metro area | 764,785 | 15,248 | 1.99% | 2.54% | 15.65% |
|  | Urban population of 2,500 to 19,999, not adjacent to a metro area | 183,613 | 3,953 | 2.15% | 0.61% | 14.84% |
|  | Completely rural or less than 2,500 urban population, not adjacent to a metro area | 235,821 | 4,904 | 2.08% | 0.78% | 16.21% |
| **Totals** |  | **30,139,792** | **544,929** | **1.81%** | **100.00%** | **15.28%** |
| 1. Derived from the Johns Hopkins University COVID-19 Dashboard from January 1, 2020, through March 31, 2021.<sup>2</sup> |  |  |  |  |  |  |
| 2. Derived from CDC Surveillance Data for Vaccinations administered at the county level through March 31, 2021.<sup>53</sup> |  |  |  |  |  |  |

## Notes

### Competing Interest Statement

The authors have declared no competing interest.

### Funding Statement

The project described was supported by the National Institute of General Medical Sciences, U54GM104942-05S2, U54GM115458, U54GM104940, U54GM104938, U54GM115516, U54GM115677, U54GM115428, and U54GM104941. The content is solely the responsibility of the authors and does not necessarily represent the official views of the NIH.

### Author Declarations

National Institute of Health's (NIH) National COVID Cohort Collaborative (N3C) Data Utilization Request Approval committee approved the data The N3C data transfer to NCATS is performed under a Johns Hopkins University Reliance Protocol # IRB00249128 or individual site agreements with NIH. The study protocol was reviewed by the N3C Data Access Committee: RP-504BA5. Use of the N3C data for this study is authorized under the following IRB Protocols: -University of Nebraska Medical Center, Omaha, Nebraska: #050-21-EP -Pennington Biomedical Research Center, Baton Rouge, Louisiana: #2021-016-PBRC -Louisiana State University Health Sciences Center, New Orleans, Louisiana: LM #1776 -West Virginia University, Morgantown, West Virginia: #2012192778 -University of Mississippi Medical Center, Jackson, Mississippi: #2020V0280 -Maine Medical Center Research Institute, Scarborough, Maine: #1697848-2 -University of Oklahoman, Norman, Oklahoma: #12947

